# Circulating H3K27 Methylated Nucleosome plasma concentration: a synergistic information with ctDNA Molecular Profiling

**DOI:** 10.1101/2023.06.02.23290872

**Authors:** Emmanuel Grolleau, Julie Candiracci, Gaelle Lescuyer, David Barthelemy, Nazim Benzerdjeb, Christine Haon, Florence Geiguer, Margaux Raffin, Nathalie Hardat, Julie Balandier, Rémi Rabeuf, Lara Chalabreysse, Anne-Sophie Wozny, Guillaume Rommelaere, Claire Rodriguez-Lafrasse, Fabien Subtil, Sébastien Couraud, Marielle Herzog, Lea Payen-Gay

## Abstract

**Background:** Molecular profiling of circulating tumor DNA (ctDNA) is a helpful tool for cancer treatment indication or for the early detection of relapse. A subset of patients with advanced lung adenocarcinoma cancers (NSCLC)can be cured by immunotherapy, radiotherapy, and/or chemotherapy combined regimens, or targeted therapies depending on their ctDNA molecular profile. However, clinical interpretation of ctDNA negative result remains challenging. Cell-free DNA (cfDNA) in association with nucleosomes are released into the bloodstream upon cell death therefore the characterization of both may provide useful information for patient management., Dysregulations of epigenetic modifications, such as histone methylation, are found to play a key role in tumorigenesis of different cancers. However, the concentration of circulating nucleosomes in blood, as a biomarker of the contributive value of ctDNA molecular profiling in patient management at diagnosis or during patient follow-up has not previously been investigated.

**Results:** Significantly elevated concentrations of H3K27Me3-nucleosomes were found in plasmas at diagnosis and during the follow-up of NSCLC patients compared to healthy donors (median: 24ng/ml; 16.9ng/ml vs 8ng/ml, p-value<0.0001, respectively). Interestingly, by combining H3K27Me3 level and ctDNA molecular profile, we found that 25.5% of the patients had high levels of H3K27Me3 (above cut-off level at 22.5 ng/ml) and no somatic alteration detected at diagnosis. This strongly supports the presence of non-mutated ctDNA in the corresponding plasma. During patient follow-up, H3K27Me3 level was lower in ctDNA-negative group compared to ctDNA-positive group (medianctDNA-= 13.4 ng/mL vs medianctDNA+ = 26.1 ng/mL, respectively, p_value<0.0001). In 41.8% of the samples, no somatic mutation and low level of H3K27Me3-nucleosomes were observed suggesting molecular indicator of treatment response. In contrast, high H3K27Me3-nucleosome level was found in 15.1% of the sample despite no somatic mutations being detected allowing the identification of disease progression from 43.1% to 58.2% over molecular profiling alone.

**Conclusion:** Measuring H3K27Me3-nucleosome levels in combination with ctDNA molecular profiling may not only improve confidence in the negative molecular result in cfDNA in lung cancer at diagnosis, it may also be a promising biomarker for Molecular Residual Disease (MRD) monitoring during and/or after treatment.

## BACKGROUND

Nucleosomes are small fragments of chromosomes released into the blood during cell death and consist of a histone octamer core with DNA wrapped around it. Interestingly, the size distribution of the DNA released into circulation (cell free DNA (cfDNA)) corresponds to nucleosomes (≍ 147bp) or chromatosome (nucleosome + DNA linker; ≍ 167bp) (1). CfDNA released into the blood following cell death exists in nucleosomal structures and encapsulated in extracellular vesicles which protects them from degradation rather as free DNA fragments (2, 3, 4, 5). In patients with cancer, cfDNA/nucleosomes are released from tumor cells and may harbor tumor DNA (ctDNA) and a variety of histone of post-translational modifications (PTMs) (6, 7).

Histone proteins PTMs such as methylation, acetylation, phosphorylation or citrullination regulate most of DNA-templated processes, including replication, transcription and repair. They function as platforms for the recruitment of specific effector proteins, such as transcriptional regulators or chromatin remodelers (8). They can also change the chromatin structure by disrupting the electrical charges in the histone residues and thus modulate accessibility to the DNA (9, 10). Histone methylation is dynamically controlled by various lysine histone demethylase (HDM) and histone methyltransferase (HMT) enzymes that remove or add the methyl group(s) from the lysine residues. Mis-regulation of the HDM and HMT leads to aberrant levels of histone methylation and have been associated with a variety of cancer types including breast, prostate, lung and brain cancers (11). For example, the HMT enzyme, Enhancer of Zeste Homolog (EZH2), the catalytic subunit of polycomb repressive complex 2 (PRC2), responsible for the trimethylation of histone H3 at lysine 27 (H3K27Me3), has been reported to be up regulated in cancer (12, 13). Its overexpression also correlates with breast cancer aggressiveness and poor prognosis (14). In parallel, H3K27Me3 levels are associated with transcriptional repression and have been reported to play an important role in the development and progression of several cancers including lung cancer (15, 16, 17). EZH2-catalyzed H3K27Me3 is also a master regulator of epithelial mesenchymal transition (EMT), an early cellular reprogramming event to facilitate the initial steps of the metastatic cascade (18, 19, 20). EZH2 levels are impaired in tumor angiogenesis and associated with drug resistance phenotype in multiple cancers (21, 22).

Epidermal growth factor receptor (EGFR)-tyrosine kinase inhibitors (TKI), such as Osimertinib and Gefitinib, have only been proven to be beneficial for patients with EGFR sensitizing genetic alterations in lung cancer (23). Recently, it had been shown that EZH2 inhibitors could sensitize *in vitro* EGFR wild-type cells to Gefitinib (24). EZH2 is highly expressed in Non-Small Cell Lung Cancer (NSCLC) and is associated with poor prognosis in patients with lung adenocarcinoma. Therefore, targeting H3K27Me3 via EZH2 is a promising cancer therapy. Tazemestostat, is the only EZH2 inhibitor approved by FDA for adults and pediatric patients aged 16 years and older. Currently, there are several ongoing clinical trials for drugs targeting EZH2 in different cancer types (25).

Combined ctDNA molecular testing to analyze circulating nucleosomes and their histone PTMs before and after patient’s treatment could provide a wider view of the genetic and the epigenetic landscape of a tumor and response to treatment. This would also enable minimal residual disease (MRD) assessment. Therefore, we conducted this retrospective, non-interventional study to i) to determine, at diagnosis, the level of circulating nucleosomes containing methylated histone residues, especially H3K27Me3-nucleosomes as potential biomarkers in patients with stage IV NSCLC; ii) to assess, during treatment, the dynamic levels of H3K27Me3-nucleosomes in NSCLC with different mutated ctDNA status to evaluate MRD to drive the choice of the following treatment line. We studied the ctDNA analysis association with nucleosome biomarkers in two independent cohorts of NSCLC patients: a first one at diagnosis before any treatment, a second one undergoing treatment including chemotherapy, combined or not with immunotherapy, or targeted therapy. Nucleosome measurements were performed on the same sample as the molecular ctDNA testing on the routine setting. This strengthens the accuracy of using biomarkers in patient management of NSCLC.

For the first time, we report a high level of circulating H3K27Me3-nucleosomes in NSCLC samples compared to healthy samples and the potential benefit of the combination of information generated by the measurement of H3K27Me3-nucleosome levels in association with the molecular ctDNA profile at diagnosis for the patient management. We also highlight the putative role of circulating H3K27Me3-nucleosomes as blood-based biomarkers for quantifying the MRD to monitor NSCLC patients during treatment.

## Results

### High levels of circulating H3K27Me3-nucleosomes are observed in NSCLC samples at diagnosis

The concentration of circulating nucleosomes with specific methylated marks H3K27Me3-, H3K36Me3-, H3K9Me3- and H3K4Me2-were measured using chemiluminescent Nu.Q^®^ immunoassays in NSCLC samples (training set (T) n_T_ = 203; validation set (V) n_V_ = 116) and compared to healthy samples (n_T_ = 100; n_V_ = 101). In both sets, we observed significantly higher circulating H3K27Me3-, H3K36Me3- or H3K9Me3-nucleosome levels in NSCLC samples compared to healthy (median_T_: 22.7 ng/mL *vs* 6.1 ng/mL; 21.8 ng/mL *vs* 11.8 ng/mL; 15.2 ng/mL *vs* 5.7 ng/mL, respectively; *p*-value < 0.0001 for each and median_V_: 28.3 ng/mL vs 9.6 ng/mL; 24.0 ng/mL vs 10.0 ng/mL; 20.4 ng/mL vs 7.5 ng/mL, respectively; *p*-value < 0.0001 for each) (Fig. 1A, B). A lower difference was observed for H3K4Me2-nucleosome levels between NSCLC and healthy samples (median_T_: 11.3 ng/mL *vs* 10.6 ng/mL and median_V_ = 12.4 ng/mL *vs* 11.2 ng/mL, respectively; *p*-value < 0.01 for each), with a very small magnitude (Table S1). The highest change observed between the medians of tumor and healthy samples was observed for the level of H3K27Me3-nucleosomes with a fold change of 3.7 and 2.9 in training and validation sets, respectively (Table S1). This marker also showed the best area under the ROC (receiving operating characteristics) curve (AUC) with a value of 0.92 (95% CI: 0.89 - 0.95) and 0.88 (95% CI: 0.84 - 0.93) corresponding to the training and validation sets analysis (Fig. 1C, D and Fig. S1A, B). The sensitivity of the H3K27Me3 assay reached 69.5% (95% CI: 61.8% - 74.5%) at 95% specificity in the training set. These clinical performances were confirmed in the validation set where the sensitivity reached 68.7% (95% IC: 59.7% - 76.5%%) at 95% specificity (Fig. 1C, D and Table S2A, B).

**Figure 1.**
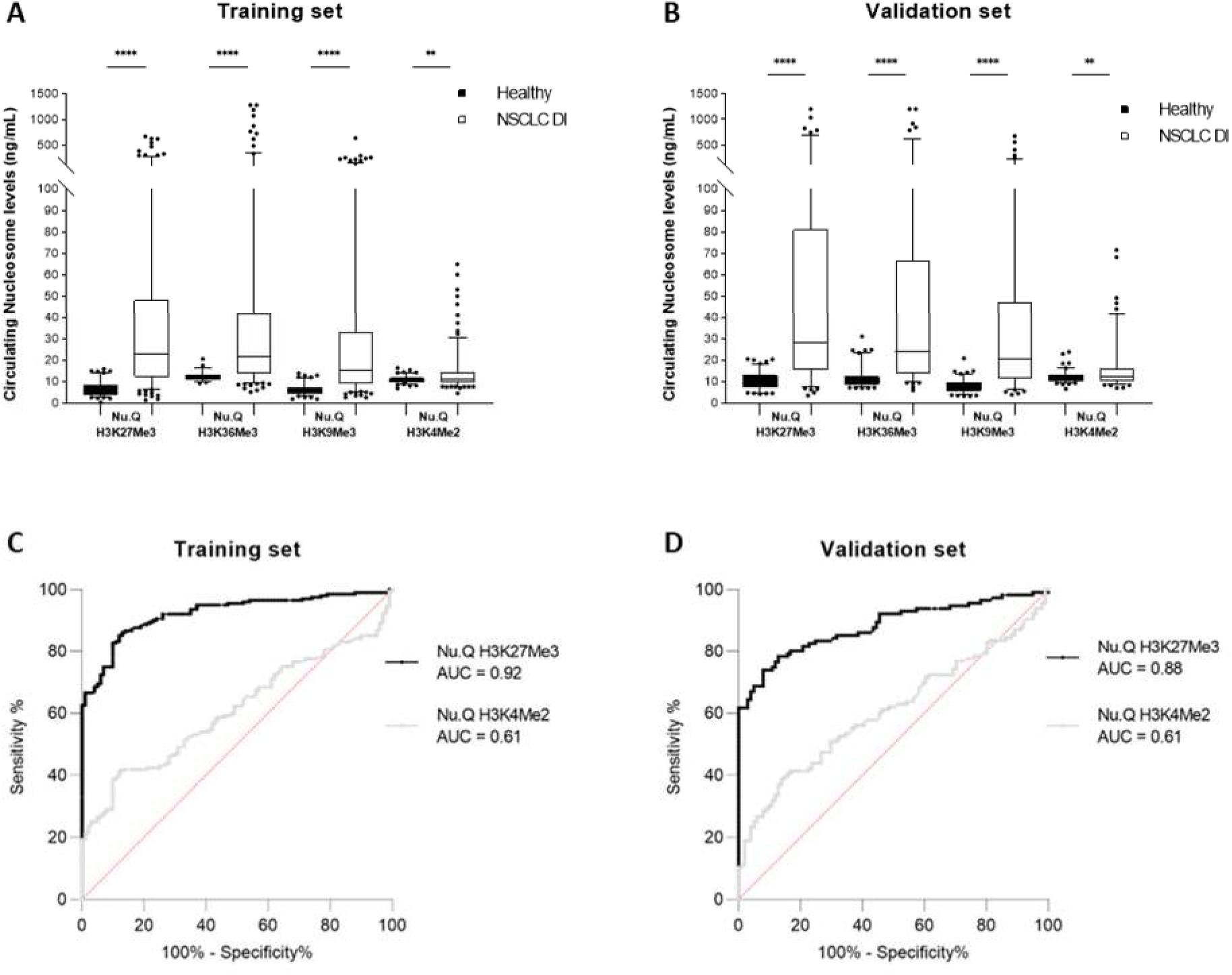
Quantification of circulating methylated-nucleosomes in NSCLC samples at diagnosis and in healthy samples. The concentration of circulating H3K27Me3-, H3K36Me3-, H3K9Me3- or H3K4Me2-nucleosomes were measured using chemiluminescent Nu.Q^®^ immunoassays on human K2-EDTA plasma samples from NSCLC patients at diagnosis (DI) and healthy donors. **A-B.** Box plot analysis representations circulating H3K27Me3-, H3K36Me3, H3K9Me3 and H3K4Me2-nucleosome levels in NSCLC samples compared to healthy samples in Training (A) and validation (B) sets. Boxes represent 25^th^-75^th^ percentile with median. Whiskers represent 2.5^th^-97.5^th^ percentile. *** and **** represent *p*-value < 0.001 and < 0.0001, respectively, calculated by Mann-Whitney U test. **C-D.** Receiver-operating characteristic (ROC) curve analysis of circulating H3K27Me3- and H3K4Me2-nucleosomes for discrimination of NSCLC at diagnosis versus healthy samples in Training (C) and validation sets (D). The areas under the curve (AUC) for the targeted nucleosome markers are indicated in legend. The red line indicates the theoretical random chance. The corresponding significance *p*-values for circulating H3K27Me3- and H3K4Me2-nucleosomes are respectively < 0.0001 and 0.0021 for the training set, *p* < 0.0001 and *p* = 0.0063 for the validation set.

### H3K27Me3-nucleosome levels correlate with cfDNA quantity and percentage of Mutational Allele Fraction (MAF) at diagnosis

Circulating nucleosomes containing the methylated histone marks and quantity of cfDNA or the percentage of mutated allele MAF correlated well (Fig. 2). The training and validation sets were combined into one whole cohort to increase the number of samples per group for better accuracy (n*_NSCLC_* = 319; n*_HEALTHY_* = 201). At diagnosis, patients with NSCLC showed a strong positive correlation between the levels of cfDNA and H3K27Me3-,H3K36Me3-, or H3K9Me3-nucleosome (*r* = 0.78; 0.66 and 0.75, respectively, *p*-value < 0.0001) and a moderate correlation with H3K4Me2-nucleosome (*r* = 0.53; *p*-value < 0.0001). The percentage of MAF for genomic somatic alterations in plasma samples was defined using the ratio absolute count of read between the reference and mutated forms. All H3K27Me3-, H3K36Me3-, H3K9Me3-,H3K4Me2-nucleosome levels and cfDNA concentration showed a weak, but statistically highly significant correlation coefficient with the percentage of MAF (*r* = 0.33; 0.31; 0.32 and 0.21, respectively; *p*-value < 0.0001 and *r* = 0.28; *p*-value < 0.001, respectively) (Fig. 2).

**Figure 2.**
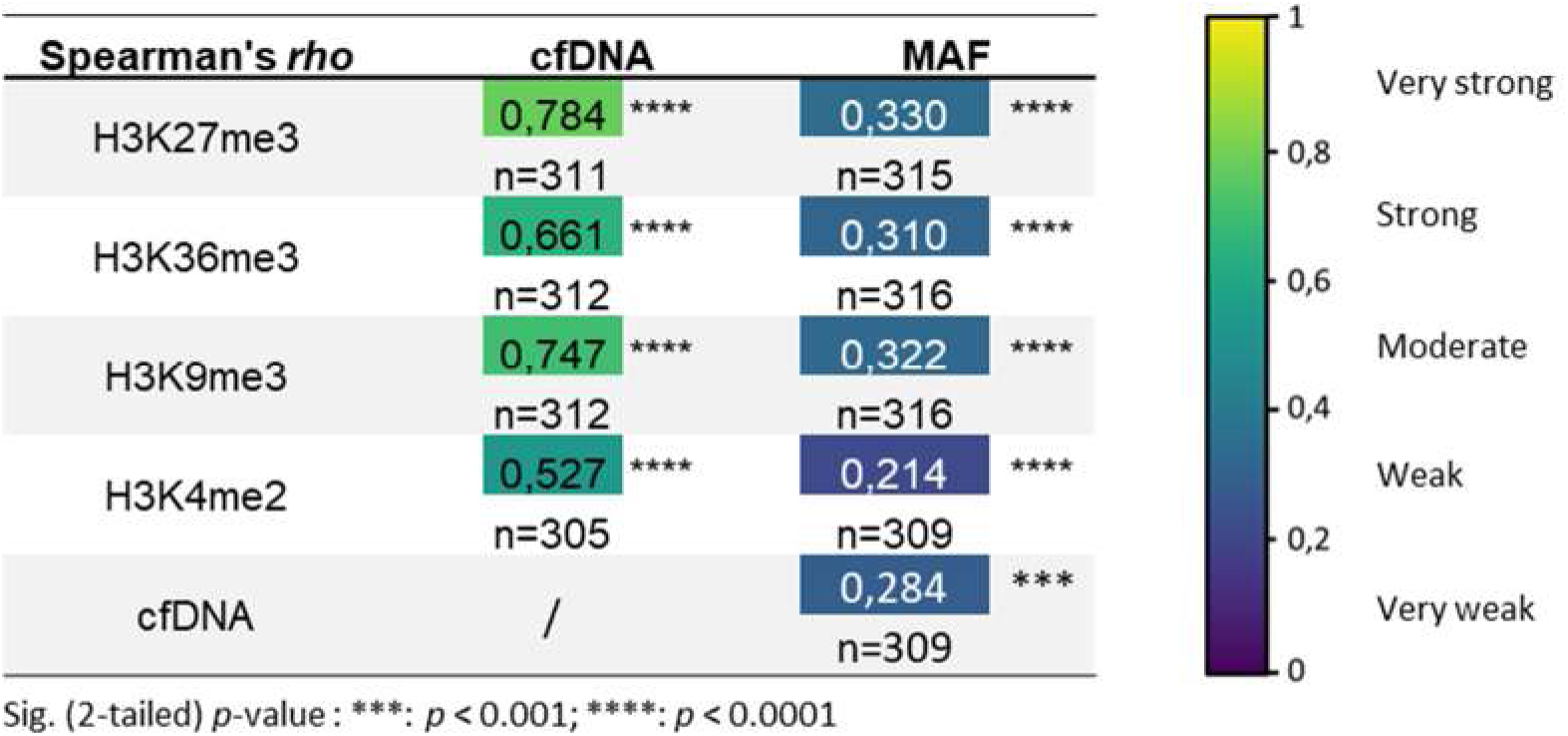
Correlation between methylated-nucleosome levels, cfDNA concentration and MAF percent in global NSCLC cohort at diagnosis. MAF % was given by SOPHIA DDM^TM^ bio-informatics analysis and corresponds to ratio of the absolute count of read between mutated forms on reference forms at each genomic position. Spearman’s rank correlation coefficient (Spearman’s rho) was calculated for combinations represented in table. *** and **** represent *p*-value < 0.001 and < 0.0001, respectively. The number of samples involved are indicated (n =) for every correlation. Spearman’s correlation coefficient ranges from 0 to 1 is represented on the right panel as a color map.

### H3K27Me3 expression level in normal and adenocarcinoma tissue

The immunohistochemical investigation of tissue microarrays (TMAs) of normal pneumocytes (n = 18) showed moderate to high H3K27Me3 expression level in 83% of cases (Fig. 3A). In the alveolar region (parenchyma) of normal lung tissue, where air and blood are brought in closed proximity over a large surface, the barrier between air and blood consists of a continuous alveolar epithelium (a mosaic of type I and type II alveolar epithelial cells), a continuous capillary endothelium and the connective tissue layer in-between, as showed in Fig. 3A. In contrast in tumoral tissue (n = 50, two cores per patient), remodeling structural events occurred (Fig. 3B, C, D). We observed i) weak H3K27Me3 expression level in 42% of cases (Fig. 3B), ii) moderate expression level in 37% of cases (Fig. 3C), and iii) high expression level in 21% of cases (Fig. 3D). TMAs of lung adenocarcinoma showed heterogeneous H3K27Me3 expression levels within the same patient demonstrating an intra-tumoral heterogeneity (data not shown). The analysis of H3K27Me3 expression as a function of grade showed that the expression of H3K27Me3 is associated with the tumor grade. Higher H3K27Me3 expression was observed in tumor grades 2 and 3 (Fig. 3E). In contrast, H3K27Me3 expression heterogeneity was not associated with the tumor grade.

**Figure 3.**
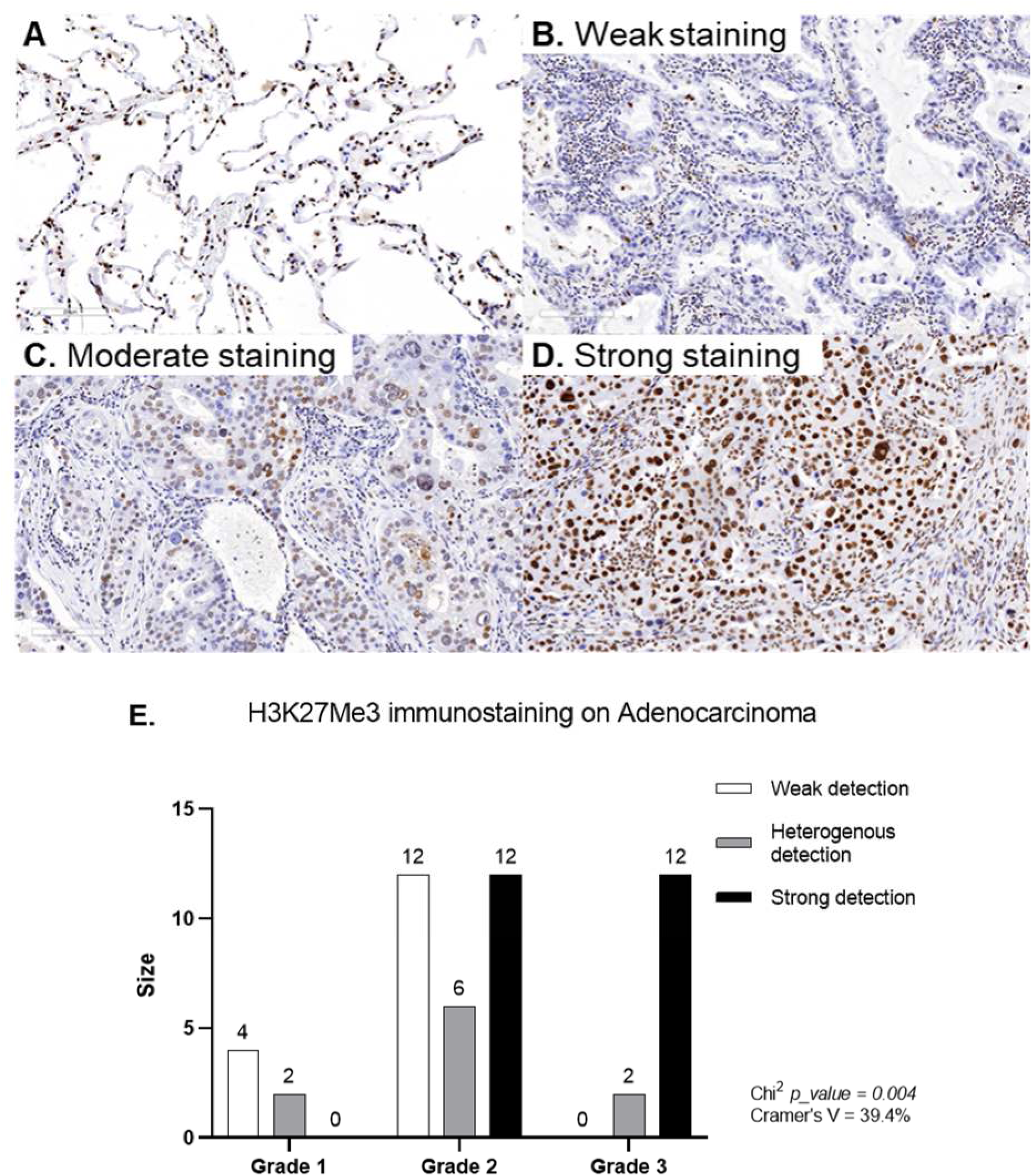
Representative H3K27Me3 immunostaining (10x objective). **(A)** representative image of normal lung tissue. **(B-D)** Representative images of weak (B), moderate (C) and high (D) intensity immunostaining from lung adenocarcinoma. **(E)** H3K27Me3 expression in function of grade.

### Relevant combined information from circulating H3K27Me3-nucleosome levels and ctDNA molecular profile at diagnosis

As H3K27Me3-nucleosome level is high in NSCLC samples but showed a low, although significant correlation, with MAF; we asked if H3K27Me3 levels could bring additional information relative to the potential presence of circulating tumor material or process. To address this question, we first defined the upper limit of the reference interval of H3K27Me3-nucleosomes in a healthy population. As the distribution of the training and validation sets were similar (Fig. S2), we combined the two sets of samples into a global and homogenous healthy cohort (n = 201) in which no outlier was identified using both the D/R ratio and ROUT methods. The upper limit of the reference interval including 95% of this population was calculated at 18.4 ng/mL (SD = 4.07). Then, we defined a cut-off at 22.5 ng/mL (18.4+1SD) ensuring 100% specificity in both training and validation sets. In the whole NSCLC cohort (n = 318), 53.1% of the patients (n = 169) showed H3K27Me3-nucleosome levels above the cut-off (Fig. 4A). The presence or absence of somatic alterations was reported on the same plasma samples based on DNAseq next-generation sequencing (NGS) analysis (the comprehensive panel contains the major 78 oncodrivers known in NSCLC). In the whole NSCLC cohort, there were 41.2% samples for which at least one somatic alteration or copy number variation (CNV) was identified, referred to hereafter as ctDNA positive sample (ctDNA+; n = 131) and 58.8% of patients for whom no genetic somatic alteration was found and defined as ctDNA negative (ctDNA-; n = 187) hereafter. The level of H3K27Me3-nucleosomes is significantly higher in ctDNA+ group compared to ctDNA-group (median = 33.9 ng/mL vs 18.5 ng/mL; *p-*value < 0.001) and compared to healthy group (median = 33.9 ng/mL vs 8.0 ng/mL; *p-*value < 0.0001) (Fig. 4B). H3K27Me3-nucleosome levels were also higher in the ctDNA-group compared to healthy group (median = 18.5 ng/mL vs 8.0 ng/mL; *p*-value < 0.0001).

**Figure 4.**
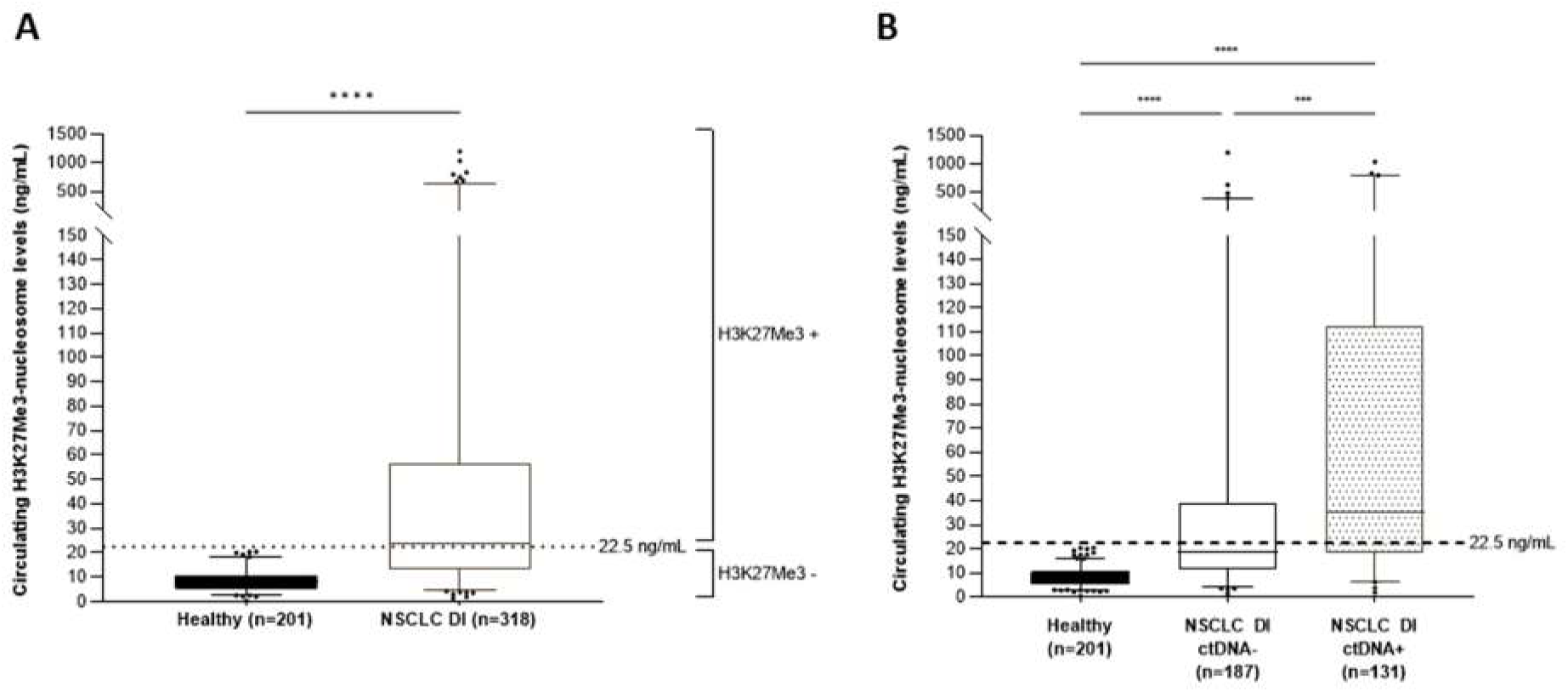
Quantification of circulating H3K27Me3-nucleosomes in NSCLC at diagnosis and in healthy samples in whole cohorts. **(A)** Healthy samples (black) and NSCLC samples at diagnosis (DI) (white). **(B)** Healthy samples (black), NSCLC samples at DI in which somatic mutation was detected (ctDNA+; dot filled) or not (ctDNA-; white), according to the somatic molecular profile obtained with a panel of 78 genes containing the major oncodrivers known and in NSCLC samples at DI with mutated ctDNA detected. The boxes represent 25^th^-75^th^ percentile with median. Whiskers represent 2.5^th^-97.5^th^ percentile. *** and **** represent *p*-value < 0.001 and < 0.0001, calculated by Mann-Whitney (groups of interest (k) = 2) and Kruskal-Wallis (*k*>2) tests.

Based on these observations, we compared the tumor mutation burden defined by the ctDNA status of the samples and the H3K27Me3-nucleosome concentrations in a decision tree model (Fig. 5). We sub-classified the samples based on their ctDNA status (ctDNA- or ctDNA+) and based on their H3K27Me3-nucleosome levels defined as H3K27Me3 positive (H3K27Me3+) or H3K27Me3 negative (H3K27Me3-) depending on whether H3K27Me3-nucleosome levels were above or below the defined cut-off at 22.5 ng/mL. We observed that: i) 33.3% of samples were double negative for ctDNA and H3K27Me3; ii) 25.5% of samples were negative for somatic alteration but had a high level of H3K27Me3-nucleosomes; iii) 13.5% samples had a low level of H3K27Me3, even if somatic alterations had been detected; iv) 27.7 % samples were found to be positive for both ctDNA and H3K27me3-nucleosomes (Fig. 5).

**Figure 5.**
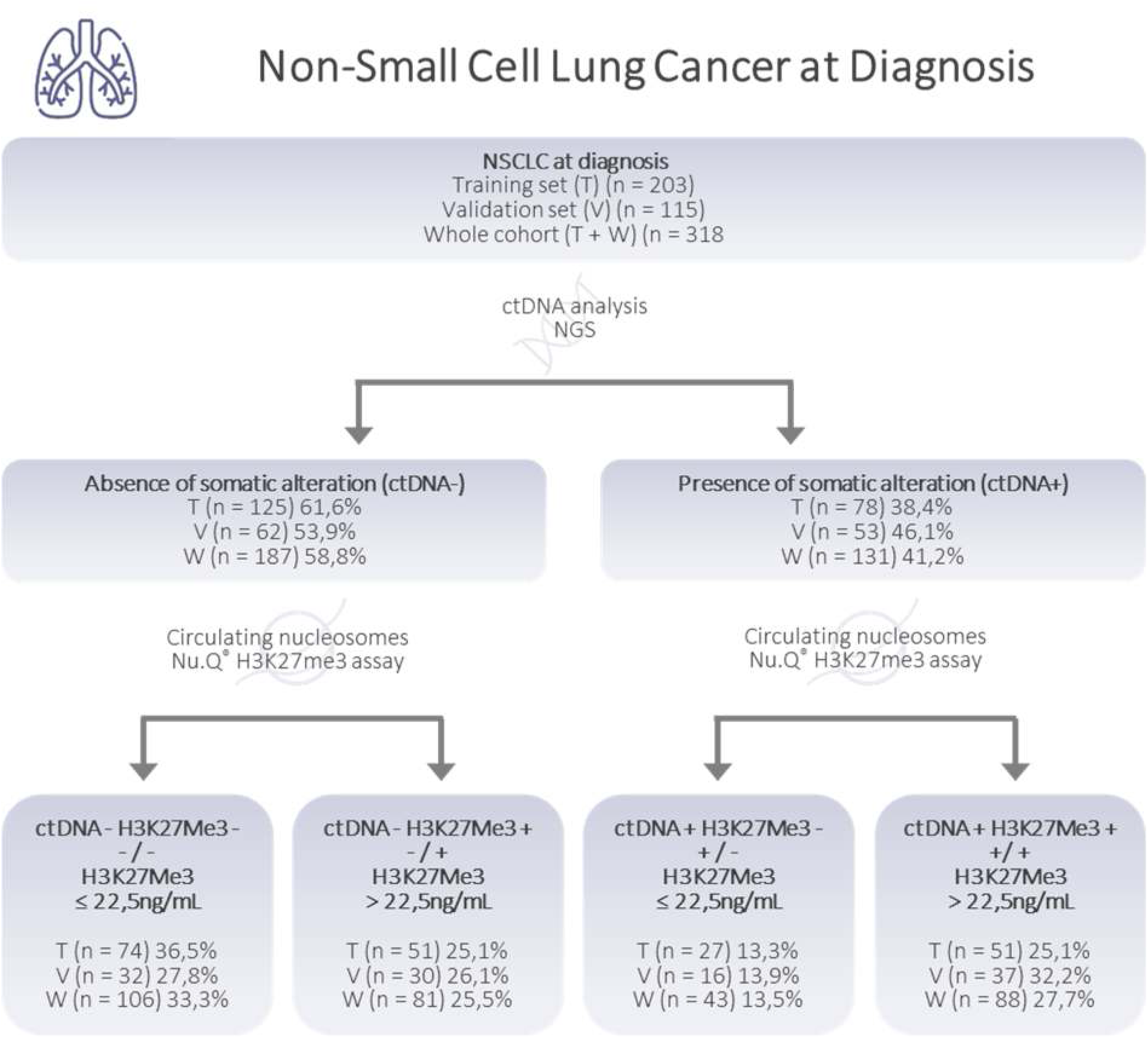
Decision tree proposed for the classification of NSCLC samples at Diagnosis. The decision is based on presence/absence of circulating tumor DNA (ctDNA) and on H3K27Me3-nucleosome levels below or above 22.5 ng/mL determined on K2-EDTA plasma samples from NSCLC patients at diagnosis. Number of samples (n =) are presented for the training (T; n = 203), the validation (V; n = 115) and the whole cohort (V+T; n = 318). Percentages express the part of the total cohort involved.

### High level of circulating H3K27Me3-nucleosomes observed in NSCLC samples during treatment is more pronounced in presence of mutated ctDNA

Based on the results observed at diagnosis, we decided to focus our analysis of NSCLC samples collected during patient treatment on H3K27Me3 only to evaluate its level change across the treatment. We assessed the concentration of circulating H3K27Me3-nucleosomes in NSCLC samples (n = 304) in routine settings and compared to healthy samples (n = 201). A highly significant increase was observed in NSCLC compared to healthy samples (median = 16.9 ng/mL vs. 8 ng/mL, respectively; *p*-value < 0.0001) (Fig. 6A). In this NSCLC population, the H3K27Me3-nucleosome levels were lower than samples collected at diagnosis (median_DURING TREATMENT_ = 16.9 ng/mL vs median_AT DIAGNOSIS_ = 24 ng/mL *p*-value < 0.0001).

**Figure 6.**
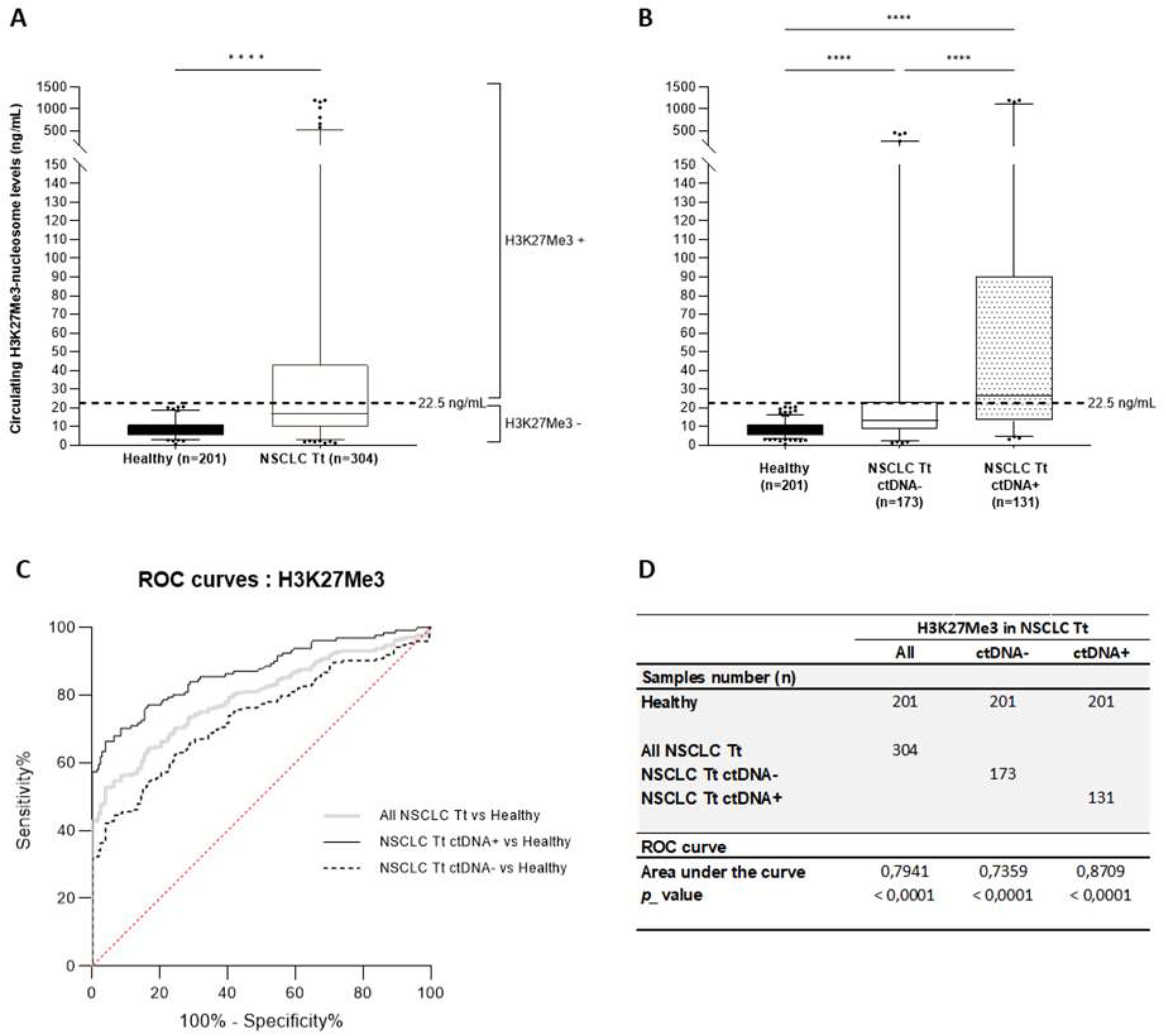
Quantification of circulating H3K27Me3-nucleosomes in NSCLC and healthy samples in the whole cohort during treatment. The concentration of circulating H3K27Me3-nucleosomes was measured using chemiluminescent Nu.Q^®^ immunoassays on human K2-EDTA plasma samples from NSCLC patients during treatment (Tt) and from healthy donors. **A-B.** The box plot analyses representing circulating H3K27Me3-nucleosome levels **(A)** in NSCLC during treatment (white) compared with healthy samples (black). **(B)** or in NSCLC samples during treatment in which mutation is detected (ctDNA+; dot filled) or not (ctDNA-; white), according to the somatic molecular profile obtained with a panel of 78 genes containing the major oncodrivers known in NSCLC. The boxes represent 25^th^-75^th^ percentile with median. Whiskers represent 2.5^th^-97.5^th^ percentile. *** and **** represent *p*-value < 0.001 and < 0.0001, calculated by Mann-Whitney (groups of interest (k) = 2) and Kruskal-Wallis (*k*>2) tests. **C.** Receiver-operating characteristic (ROC) curve analysis of circulating H3K27Me3-nucleosomes. Discrimination of NSCLC during treatment versus healthy groups (grey line) is compared to discrimination of NSCLC ctDNA+ during treatment versus healthy groups (black line) and to discrimination of NSCLC ctDNA-during treatment versus healthy groups (dotted line). The red line indicates the theoretical random chance. **D.** The groups size, areas under the curve (AUC) and corresponding *p*-values for the targeted groups are listed. All analyses are highly significant.

The molecular profiles of these NSCLC samples were conducted by NGS analyses. In 43.1% of the samples (n =131), at least one mutation among the 78 screened genes was detected, demonstrating the presence of ctDNA in plasma samples. Then, we compared the circulating H3K27Me3-nucleosome levels in positive and negative ctDNA samples (ctDNA+, ctDNA-) collected during the treatment. The level of circulating H3K27Me3-nucleosomes was lower in the ctDNA-negative group compared to ctDNA-positive group (median_ctDNA-_ = 13.4 ng/mL vs median_ctDNA+_ = 26.1 ng/mL, respectively, *p*-value < 0.0001) (Fig. 6B). A greater heterogeneity of H3K27Me3-nucleosome levels was observed in presence of somatic alteration, 55% of samples were found above the H3K27Me3-nucleosomes cut-off in the ctDNA+ group, whereas only 26.6% of the samples were above in the ctDNA-group (Fig. 6B). The clinical performance was then evaluated on the whole cohort (in grey), on the ctDNA-(dotted line) and ctDNA+ (in black) sub-groups in comparison to the healthy samples (Fig. 6C, Table S3). We found that the discrimination of NSCLC samples was improved in ctDNA+ sub-group with an AUC of 0.87 compared to 0.74 for ctDNA-sub-group and 0.79 for the whole cohort respectively; *p*-value < 0.0001) (Fig. 6C, D).

### Additive value of both biomarkers: H3K27Me3-nucleosomes and ctDNA

After sub-grouping of NSCLC samples collected during treatment into ctDNA- and ctDNA+ classification; samples were further classified based on their level of circulating H3K27Me3-nucleosomes below or above the cut-off at 22.5 ng/mL, as H3K27Me3-negative (H3K27Me3-) or H3K27Me3-positive (H3K27Me3+) and organized in four classes described in a decision tree (Fig. 7A). We observed i) 41.8% of samples are double negative for the ctDNA and circulating H3K27Me3-nucleosome levels (ctDNA-H3K27Me3-); ii) 15.1% of samples are negative for somatic alteration but positive for circulating H3K27Me3-nucleosomes (ctDNA-H3K27Me3+); iii) 19.4% of samples had a level of circulating H3K27Me3-nucleosomes below 22.5 ng/mL, even when somatic alterations had been detected (ctDNA+ H3K27Me3-); iv) 23.7% of samples were found to be positive for both ctDNA and circulating H3K27Me3-nucleosome levels (ctDNA+ H3K27Me3+).

**Figure 7.**
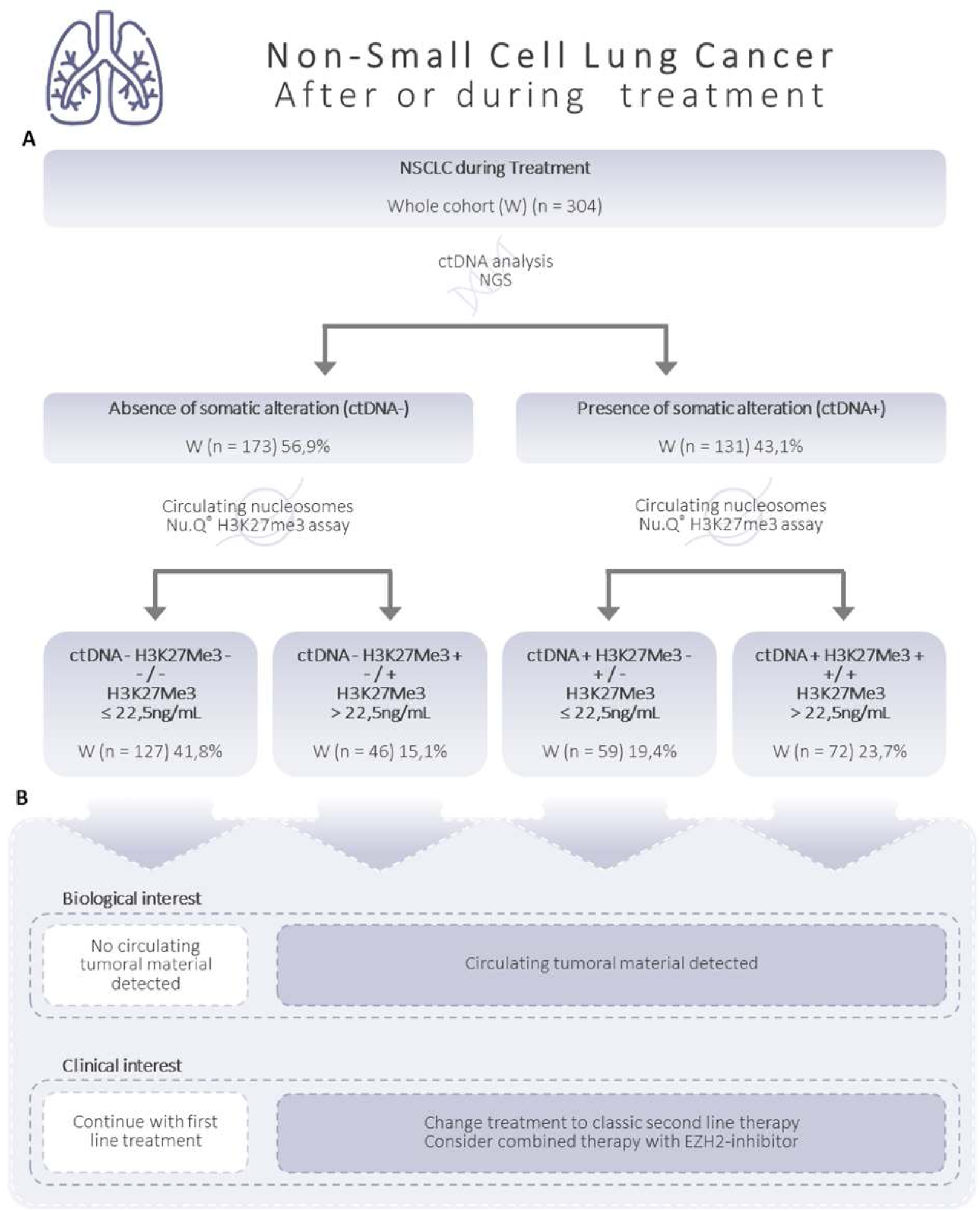
Decision tree proposed for the classification of NSCLC samples during patient follow-up. **A.** The decision is based on H3K27Me3-nucleosomes levels below or above 22.5 ng/mL and in presence/absence of circulating tumor DNA (ctDNA) determined on K2-EDTA plasma samples from NSCLC patients during treatment. Number of samples (n =) are presented for the whole cohort (W; n = 304). Percentages express the part of the total cohort involved. **B.** Biological and clinical interests are based on our proper interpretation and should be validated clinically.

### High circulating H3K27Me3-nucleosome levels are preferentially associated with TP53 mutations during treatment

In ctDNA+ group, levels of circulating H3K27Me3-nucleosomes were compared based on the mutation status of three major oncodrivers of interest: EGFR, KRAS and TP53. Mutational status in the corresponding sub-groups was annotated as positive (+) when at least one mutation was detected and negative (-) when no mutation was detected (Fig 7). In parallel of the tendency of association found at diagnosis (data not shown), significant increase of circulating H3K27Me3-nucleosome levels was observed in TP53+ (n = 72) sub-group compared to TP53-(n = 59) sub-group (median_TP53+_ = 36.9 ng/mL vs median_TP53-_ = 19.2 ng/mL; *p-*value < 0.05) (Fig. 8 A-B). No difference was observed in comparisons between EGFR+ versus EGFR-neither KRAS+ versus KRAS-sub-groups (median_EGFR+_ = 21.8 ng/mL vs median_EGFR-_ = 36.3 ng/mL; *p-* value = 0.22; median_KRAS+_ = 14.8 ng/mL vs median_KRAS-_ = 26.7 ng/mL; *p-*value = 0.50) (Fig. 8 A-B).

**Figure 8.**
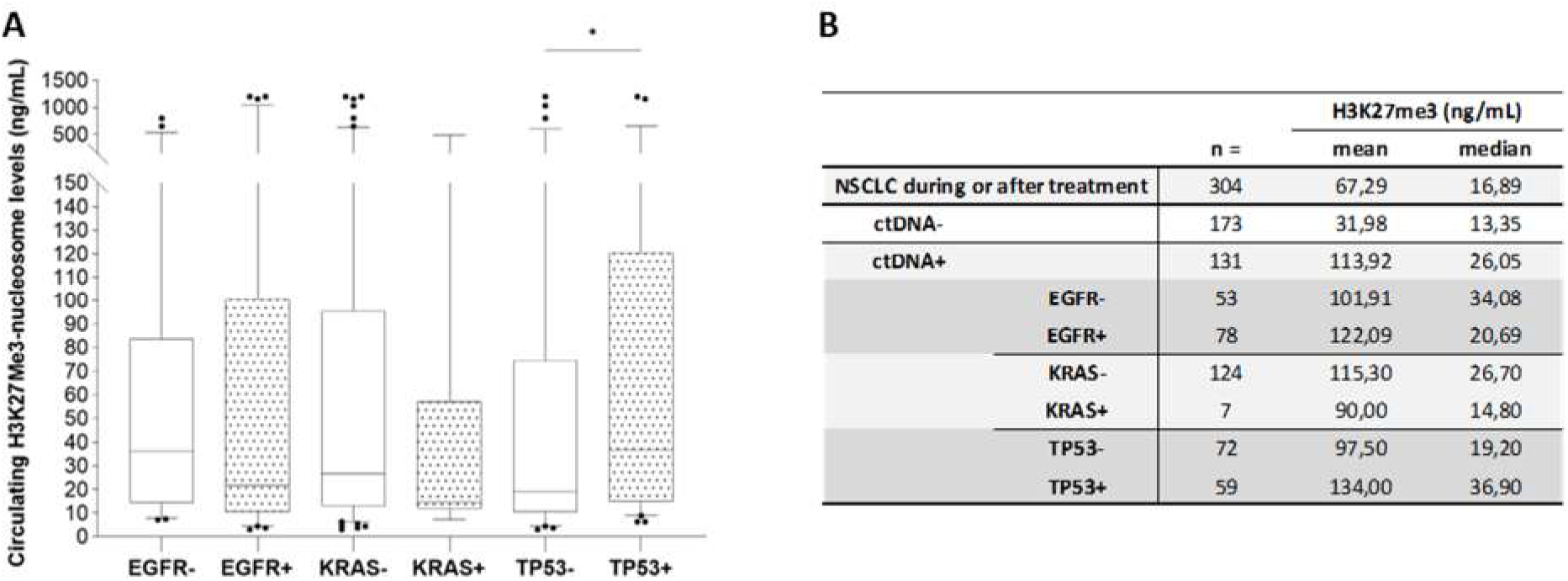
H3K27Me3-nucleosome levels of NSCLC samples during treatment according to the somatic alteration status. The concentration of circulating H3K27Me3-nucleosomes was measured using chemiluminescent Nu.Q^®^ immunoassays on human EDTA plasma samples. Only ctDNA+ group was included in this analysis. Minus (-) groups referred to samples where a mutation in an oncodriver is different from the gene of interest (*EGFR*, *KRAS* or *TP53*); plus (+) groups referred to samples for which at least one mutation is detected in the gene of interest (*EGFR*, *KRAS* or *TP53*). **A.** Boxes represent 25^th^-75^th^ percentile with median. Whiskers represent 2.5^th^-97.5^th^ percentile. * represent *p*-value < 0.05 calculated by Mann-Whitney U test for testing the hypotheses of equality or difference between the two groups of interest. **B**. Groups size (n =) means and medians are summarized in table.

## Discussion

Patient management with NSCLC is based on histopathological evaluation completed with protein and molecular biomarkers for determining the optimal treatment. Tumor tissue-based molecular testing has demonstrated its utility to assess patients for the recommended biomarkers but remains challenging due to tissue invasive sampling, DNA quantity availability and heterogeneity, and inadequacy of repeated testing (26). In contrast, genomic somatic testing of oncodrivers through liquid biopsy has emerged as a key strategy for managing patients that are diagnosed with an advanced NSCLC and can be used both at diagnosis to define first-line therapy and throughout patient follow-up to identify the emergence of resistant mechanisms. Therefore, although tissue biopsy remains the reference standard for cancer diagnosis and patient management, when combined with plasma-based molecular profiling, the patient standard of care is improved by approximately 11 to 20% (27, 28, 29). In contrast to tissue biopsies, plasma testing can be easily repeated serially to monitor response and may allow detection of minimal residual disease prior to ct-scan or clinical progression. It also reduces invasiveness and costs. In consequence, it is more compliant for patients. Despite the high clinical specificity of ctDNA, physicians often face a high non-contributive detection rate of preoperative mutated plasma ctDNA, around 24% to 85% in the overall population of lung cancers associated with stage in routine settings (30, 31, 32). CtDNA was detected after treatment in 64.3% of patients who had obvious clinical recurrence (33). Moreover, a study conducted by Chaudhuri et al., showed that disease recurrence in ctDNA negative population happened in 6% of the post-operative population of NSCLC patients (34). So currently, the interpretation of an undetectable ctDNA is still a significant limitation in clinical indication. Other circulating biomarkers may complement ctDNA molecular testing. Circulating nucleosome as the carrier of genetic and epigenetic information and genetic aberrations could be one of these. In addition, thanks to the low volumes required for the nucleosome Nu.Q^®^ immunoassay, it is easy to add it at the plasma molecular profiling testing on the same plasma sample. In this study, we focused on circulating H3K27Me3-nucleosome levels in NSCLC patients at diagnosis or during their treatment, and the combined contributive information to molecular profiling of circulating H3K27Me3-nucleosomes.

TMA analysis showed that H3K27Me3 was detected in the nucleus of normal adjacent cells and adenocarcinoma cells. Histone modification is a dynamic process as demonstrated recently by FRET (fluorescence resonance energy transfer) biosensor-based technique in cancer living cells (35). Unlike H3K9Me3 that is always associated with silenced chromatin regions, H3K27Me3 mark is found in bivalent domains associated with gene repression but still allow these genes to be activated (36). The repression was not randomly distributed within the genome but enriched in inactive topologically associated domains (TADs) with low expression of genes involved in proliferation (37, 38). As described for genetic clonality profiling and PD-L1 (programmed death ligand 1) tissue expression, we observed intra- and inter-tumoral heterogeneity in adenocarcinomas (39). Interestingly, the frequency of high expression of H3K27Me3 was larger in grade 3 adenocarcinoma compared to grade 2 and grade 1 suggesting that in NSCLC, like in thyroid cancer or invasive phenotype of melanoma, H3K27Me3 overexpression could be positively associated with tumor aggressiveness (40, 41). Cellular plasticity and tumor cells dedifferentiation is commonly observed in various types of malignancies and is described to be associated with increased tumor cell invasiveness and drug resistance. We can speculate that the association between H3K27Me3 level and tumor aggressiveness could be linked with its role in this loss of differentiation of tumor cells (42, 43, 44). At diagnosis, circulating H3K27Me3-nucleosome levels are significantly higher in NSCLC patients compared to healthy donors. In contrast to tissue observation, this analysis of circulating H3K27Me3-nucleosomes reflects the release of nucleosomes into the blood following cell death and high cell turnover occurring in cancer (45, 46, 47).

To discuss the advantage of the quantification of H3K27Me3-nucleosomes in clinical settings, we had to establish the normality range into healthy population using similar methodology as other tumoral biomarkers such as CYFRA21.1 and CA125. For doing that, we first determined the upper limit of the reference interval of H3K27Me3-nucleosomes in a healthy population and determined the threshold of positivity at 22.5 ng/mL ensuring 100% specificity in both training and validation sets. Increased H3K27Me3-nucleosome levels observed in NSCLC patients was even more pronounced in patients who have somatic alterations and was logically positively associated to the cfDNA concentration as its carrier. Somatic mutations were found in 41.2% of analyzed samples. 27.7% of the samples were positive for both ctDNA and circulating H3K27Me3-nucleosomes pointing to putative epigenetic and mutational burden processes, while in 13.5% samples only somatic alterations were detected demonstrating the presence of tumoral material in blood but a low level of H3K27Me3-nucleosome which might suggest a low-grade tumor or epigenetic changes compared to normal cells as supported by the TMA analysis performed.

Besides, one fourth of patients (25.5%) had a high H3K27Me3-nucleosome levels even if no somatic alteration were detected in plasma. This is indicative of tumoral DNA release into blood despite absence of somatic alteration detected, indicating somatic mutation not covered by the comprehensive panel of genes used in the customized NGS assay and potentially a high-grade cancer. We can hypothesize that most likely nucleosome and its associated DNA came from cancer cells but from a genomic region that did not have a cancerous mutation. Those samples could be considered as true ctDNA negative for the main oncodriver alterations. In the opposite, 33.3% of the samples were negative for both ctDNA and H3K27Me3-nucleosome level indicating no or very few tumoral material released into blood. In this case, a re-biopsy will be probably required for those patients at diagnosis until additional cancer biomarkers are identified. Altogether, these results at diagnosis showed that by combining circulating H3K27Me3-nucleosome levels with genomic testing, we can increase the contributing information of ctDNA sequencing by 25.5% to help interpret negative ctDNA results. In addition, high level of H3K27Me3-nucleosomes in half of the cases (52%) could be used as a biomarker to monitor patients during follow-up.

During patient’s treatment, high levels of circulating H3K27Me3-nucleosomes were still observed in NSCLC samples especially in presence of mutated ctDNA. Nevertheless, we can note that despite being still high, the H3K27Me3-nucleosome levels in the ctDNA negative group were closer to the one observed in the healthy group with 61.2% of the samples below the cut-off. As the study was conducted on two independent cohorts at diagnosis and during treatment, it was not possible to make a direct comparison. But Holdenrieder and al. described that in patients with advanced lung cancers, levels of circulating nucleosomes at diagnosis were significantly lower in patients who responded to chemotherapy, suggesting that nucleosome could be a potential predictive biomarker (48). When combining both ctDNA sequencing and H3K27Me3-nucleosome measurements, we observed nearly 42% of the samples negative both ctDNA and H3K27Me3-nucleosomes suggesting no detection of tumoral material and a potential positive evolution of the disease. First-line treatment should be continued, and the next follow-up may be spaced out in time (Fig. 7B). In 15% of the samples, no somatic mutation was found but a high level of circulating H3K27Me3-nucleosomes was detected. This could illustrate a progression of the disease in absence of acquisition of somatic mutation, or no alteration covered by the NGS comprehensive gene panel. A change in the treatment to classic second line therapy may be considered (Fig. 7B). These results could also suggest that a combined therapy with EZH2-inhibitor might be appropriate. More clinical data on EZH2 inhibitor performances in NSCLC are required to confirm or not this option. Somatic mutations have been detected in the remaining 43% of the samples associated with a high level of H3K27Me3-nucleosomes (23.7%) or not (19.4%) demonstrating a progression of the cancer and the need to change the treatment (Fig. 7B). Importantly, when the level of H3K27Me3-nucleosomes is considered with ctDNA there was an increased identification of disease progression from 42 to 58% of patients. In addition to the potential benefit to combine H3K27Me3-nucleosomes and ctDNA analysis at diagnosis to ascertain a true negative ctDNA results, we propose that H3K27Me3-nucleosomes could also be a good potential biomarker to detect minimal residual disease and to monitor response during disease progression. Further studies would be required to validate the potential of circulating H3K27Me3-nucleosomes as biomarker for NSCLC patient follow-up. When validated, we could consider implementing the measurement of H3K27Me3-nucleosomes before molecular profiling analysis for an improved patient care and a more cost-efficient health care system. Indeed, if during post-treatment, the patient is well with normal scan images and low H3K27Me3-nucleosomes levels, ctDNA analysis could be postponed. In contrast, high level of H3K27Me3-nucleosomes levels should trigger a quicker ctDNA analysis and therapeutic decision.

In French population, the frequency of tissue somatic alterations reported in patient with NSCLC is 11% in *EGFR*, 1% in *HER2*, 29% in *KRAS* 2% in *BRAF*, and 2% *PIK3CA* (*49*). In our study, the molecular status was not fully associated with the H3K27Me3-nucleosome concentrations. Nevertheless, the TP53 status, especially during progression, seems to be correlated with the H3K27Me3-nucleosome concentration. 40% TP53 somatic alterations are reported in NSCL (50, 51, 52). This detection is an indicator of a poor prognosis in patients with NSCLC. In advanced NSCLC patients treated with nivolumab, with or without CTLA-4 blocker ipilimumab, or pembrolizumab, the median overall survival in the TP53-mutated group was 18.1 months vs 8.1 in the TP53-wild-type group. This strongly indicated that TP53 status might be potential predictor of immunotherapy response (53). These TP53 mutations may differ according to different pathological types and clinical stages and are often detectable during progression or in aggressive grade. These findings agreed with the higher level of H3K27Me3-nucleosome into TP53 mutated samples observed in our cohorts. In this report, the H3K27Me3-nucleosome concentration was not statistically different according to the EGFR and KRAS status and must be addressed in a wider cohort.

## CONCLUSIONS

Using a simple and low-cost H3K27Me3-nucleosomes immunoassay to complete the molecular exploration of ctDNA could greatly improve confidence in the negative molecular results at diagnosis. This may allow to reduce re-biopsy invasive acts. High levels of H3K27Me3-nucleosomes could allow physicians to detect MRD in NSCLC patients at defined intervals of treatment and recovery to incorporate analysis of evolving molecular landscapes during treatment. These data should be confirmed in a longitudinal study where NSCLC patients will be followed from diagnostics to recovery or relapse.

## Methods

### Study Population

Patients were recruited from the pulmonology department at Lyon University Hospital from 2015 to 2022. K2-EDTA plasma heathy samples were provided by the French agency named “Etablissement Français du Sang” (Etude RNIPH 22-5065 NUCLEO_CIRCAN avis CSE n°22-5065). Inclusion criteria were: age over 18 years old with histologically proven NSCLC cancer. For each sample, medical data were collected through a mandatory prescription sheet attached to each sample and edited by the prescribing physician. K2-EDTA plasma samples from 319 patients at diagnosis and from 309 independent patients under treatment, defined by a clinical or CT-scan modification during their treatment were analyzed.

### CfDNA Collection

Total EDTA blood samples were centrifuged for 10 min at 1600 g. The supernatant was then centrifuged at 6000 g for 10 min, and the resulting plasma was stored at −80 °C until cfDNA extraction and molecular analyses (54). CfDNA was extracted using the QIAamp Circulating Nucleic Acid Kit (Qiagen, Valencia, CA, USA, Cat No 55114), with a Qiagen vacuum manifold following the manufacturers’ instructions. CfDNA samples were quantified using a Qubit™ 4 Fluorometer (Invitrogen™, Cat No Q33238, Carlsbad CA, 92008, United States) with the Qubit™ dsDNA HS Assay Kit (Invitrogen™, Cat No 32854).

### Library Preparation for DNA Sequencing

For custom-validated NGS library preparation, 10 – 100 ng cfDNA were used, using a custom capture-based technology provided by SOPHiA GENECTICS (Lausanne, Switzerland) and performed according to the manufacturer’s instructions (55, 56, 57). The custom panel covered 78 genes involved in cancer (such as *EGFR*, *TP53* or *KRAS*). The libraries were sequenced on NextSeq 550 (Illumina technology, San Diego, CA 92122) in 2 × 150 paired-end runs. The subsequent Variant Call Files were subjected to cross-sample background filtering, with potential artefacts removed below 3 standard deviations of the mean background noise for each position. The bioinformatics was performed using the SOPHIA DDM^TM^ platform.

### Nu.Q^®^**^®^**ImmunoAssays

Four nucleosome structures were measured using Nu.Q^®^ prototype Immunoassays: Nu.Q^®^ H3K27Me3, Nu.Q^®^ H3K36Me3, Nu.Q^®^ H3K9Me3, Nu.Q^®^ H3K4Me2 (Belgian Volition SRL, Isnes, Belgium) according to the manufacturer’s instruction. Briefly, these sandwich immunoassays are based on magnetic beads and chemiluminescence technology and are performed on the IDS-i10 automated immunoanalyzer system (Immunodiagnostic Systems Ltd (IDS), UK). 50 μL of K2-EDTA plasma (same as for the DNA sequencing) are incubated with acridinium ester labeled anti-nucleosome antibody. Then, magnetic particle beads, coated with the corresponding monoclonal anti-histone modification capture antibody (i.e., anti-histone H3K27Me3, anti-histone H3K36Me3, anti-histone H3K9Me3, or anti-histone H3K4Me2, respectively), are added. Finally, after a wash step, trigger solutions are added, and the light emitted by the acridinium ester is measured by the luminometer system. The results are expressed in relative light unit (RLU) and the concentrations are extrapolated using a four-parameter logistic regression of a reference standard curve. All samples are analyzed in simplicate.

### Tissue microarray (TMA)

Two TMAs were purchased from the company TissueArray.com. The first TMA corresponds to 100 cores of 50 cases (2 cores by case) of lung adenocarcinoma (BC04022a), while the second TMA contains 18 cases of squamous cell carcinoma, matched with adjacent normal lung tissue and cancer adjacent tissue per case (LC10014a).

The immunohistochemical analysis was performed with an automated immunostainer (BOND). Briefly, formalin-fixed paraffin-embedded 4-μm-thick sections were first19 paraffinized in xylene and rehydrated in ethanol. The endogenous peroxidase activity was blocked (Ventana Medical Systems, Tucson, Arizona, USA) before antigen retrieval was begun. The ULTRA Cell Conditioning Solution (ULTRA CC1) from Ventana Medical Systems was then used for antigen retrieval. Immunohistochemical staining was carried out on an automated immunostainer, with primary antibody. This was followed by application of the avidin–biotin– peroxidase complex technique. Reactions were developed with diamino-3,3′-benzidine tetrahydrochloride substrate solution (SIGMAFAST; Sigma-Aldrich, Tucson, Arizona, USA). The tissues were counterstained with hematoxylin. The primary antibodies and final dilutions were: H3K27Me3 (Cell Signaling Technology, Danvers, Massachusetts, USA).

Immunostaining in the tumor tissue was determined by visual scoring of the brown stain. Scoring of the intensity of the staining was performed according to an arbitrary scale with steps of 0, 1, 2, and 3 where “0” was considered to be absence of staining, “1” considered weak staining, “2” was considered as moderately positive staining, and “3” was considered to be strong staining.

### Statistics

Statistical analyses were performed using GraphPad Prism (GraphPad Prism software version 9.5.0, San Diego, CA, USA). Descriptive statistics were used, and results are reported as mean, median, 2.5^th^-25^th^-75^th^ and 97.5^th^ percentiles. Data were subjected to Kolmogorov–Smirnov normality test. As data are not normally distributed, non-parametric methods were used: Mann-Whitney U test and Kruskal-Wallis H test for groups comparison and Spearman’s rank correlation for measuring the dependence between variables. Significance values are represented by *: *p-*value < 0.05; **: *p* < 0.01; ***: *p* < 0.001; ****: *p* < 0.0001. Reference interval (also referred as normal range) of H3K27Me3-nucleosome levels was calculated on global healthy cohort to reach the statistically significant minimum number (n ≤ 120). The data were subjected to D/R ratio and ROUT methods for the detection of outliers. Reference interval includes 95 % of the population, centred on the median. One standard deviation was added to the upper limit of this reference interval to determine the cut-off.

### Study limitations

As this is a retrospective study carried out for routine use, we are facing limitations, including: i) access to clinical data is limited to ctDNA molecular profile and time of sampling in the routine patient management; ii) we did not have matched tissue-plasma samples, the histological analyses were performed with the commercial Tissue Macro-Array. iii) independent cohorts were analyzed at diagnostic and during treatment.

## DECLARATIONS

### Ethics approval and consent to participate

Patients were recruited from the pulmonology department at Lyon University Hospital from 2015 to 2022 (Etude RNIPH 22-5065 NUCLEO_CIRCAN avis CSE n°22-5065).

### Consent for publication

Medical data were collected through a mandatory prescription sheet and edited by the prescribing physician. The patients were orally informed of the testing by physicians conducting their clinical management.

### Availability of data and materials

The data presented in this study are available in the Additional Files and from the corresponding author

### Authors’ contributions

Conceptualization: E.G., D.B., M.H. and L.P.-G.; data curation: J.C., G.L., F.G., J.B. and L.P.-G., formal analysis: E.G., J.C., D.B., M.H. and L.P.-G.; funding acquisition: S.C, and L.P.-G; investigation: C.B., D.B., E.G., A.G., R.R and L.P.-G; methodology: F.S., D.B., G.L., N.H., M.H. and L.P.-G; project administration: C.R.-L., S.C., M.H. and L.P.-G.; resources: S.C. and E.G.; supervision: M.H. and L.P-G.; validation: all authors; writing—original draft: E.G., J.C., B.B., G.L., D.B., M.H. and L.P.-G; writing-review and editing: J.C., B.B., G.L., D.B., A.-S.W., S.C., M.H. and L.P.-G. All authors have read and agreed to the published version of the manuscript.

## Acknowledgements

The authors are very grateful to AstraZeneca for their long-term collaboration with CIRCAN team from Lyon-Sud Hospital. The authors thank AstraZeneca and Sysmex Inostics for their continued financial support of this clinical study. The authors wish thanks the Assay Development team from Belgian Volition for their technical support. We also thank Terry Kelly for critical reading of the manuscript.

## ADDITIONAL FILES

**Fig. S1. A-B. ROC curve analyses of circulating H3K36Me3- and H3K9Me3-nucleosomes for discrimination of NSCLC versus healthy groups.** The areas under the curve (AUC) for the targeted nucleosome markers are indicated in legend. The red line indicates the theoretical random chance. All curves were highly significantly different from the chance for circulating H3K36Me3- and H3K9Me3-nucleosomes and the *p-*value were all < 0.0001 for the training and validation sets.

Additional Figures.docx

**Fig. S2. Frequencies distribution of H3K27Me3-nucleosome levels of healthy samples in training and validation sets.** Training and validation sets are represented in white and grey bars, respectively. *y* axis represents the relative frequency of samples expressed in percent of the total cohort along the concentration of H3K27Me3 expressed in ng/mL on the *x* axis.

Additional Figures.docx

**Table S1. Fold change between levels of methylated-nucleosomes in NSCLC at diagnosis compared to healthy samples.** The fold change represents a ratio which is computed as the median level of specific circulating nucleosomes measured on NSCLC samples divided by the median level of the reference Healthy samples.

Table S1.xlsx

**Table S2. A-B. Sensitivity and specificity data for the ROC curves in Figure 1 and 6**. Data corresponding to Figures 1 C and S1. A are presented in table S2 A (Training set); data corresponding to Figures D and S1. B are presented in table S2 B (Validation set).

Table S2.xlsx

**Table S3. Sensitivity and specificity data for the ROC curve in Figure 6C**.

Table S3.xlsx

