## Supplementary figures and images for "Circulating H3K27 Methylated Nucleosome plasma concentration: a synergistic information with ctDNA Molecular Profiling"

### ADDITIONAL FILES

**ADDITIONAL FIGURES**

**Fig. S1. A-B.**

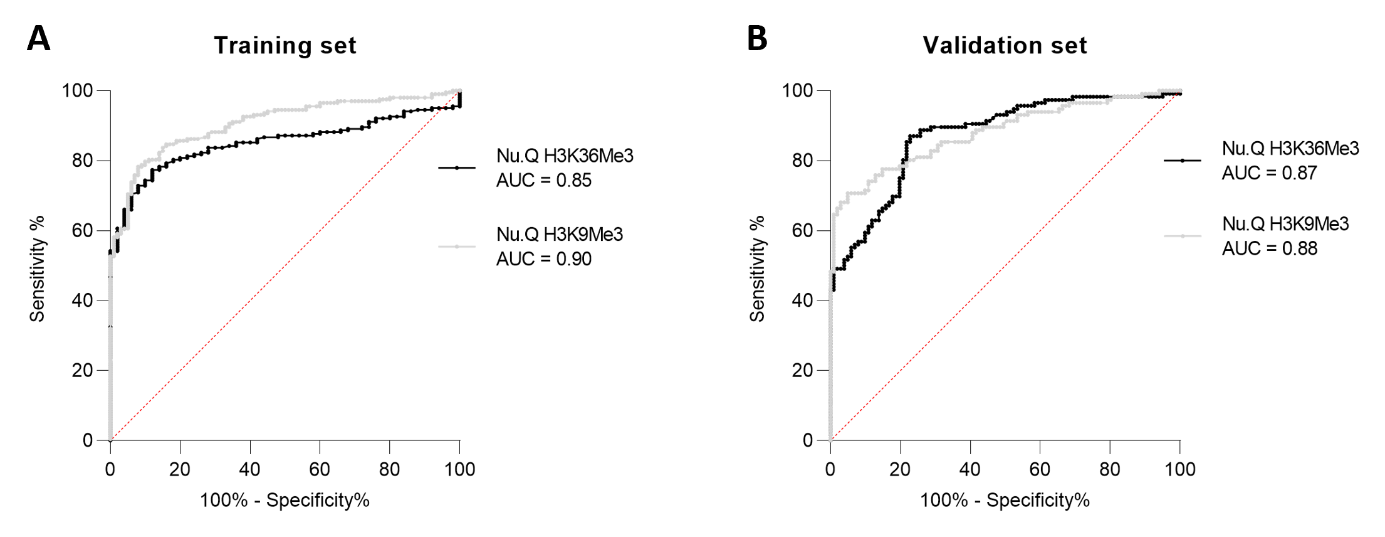

**Fig. S2.**

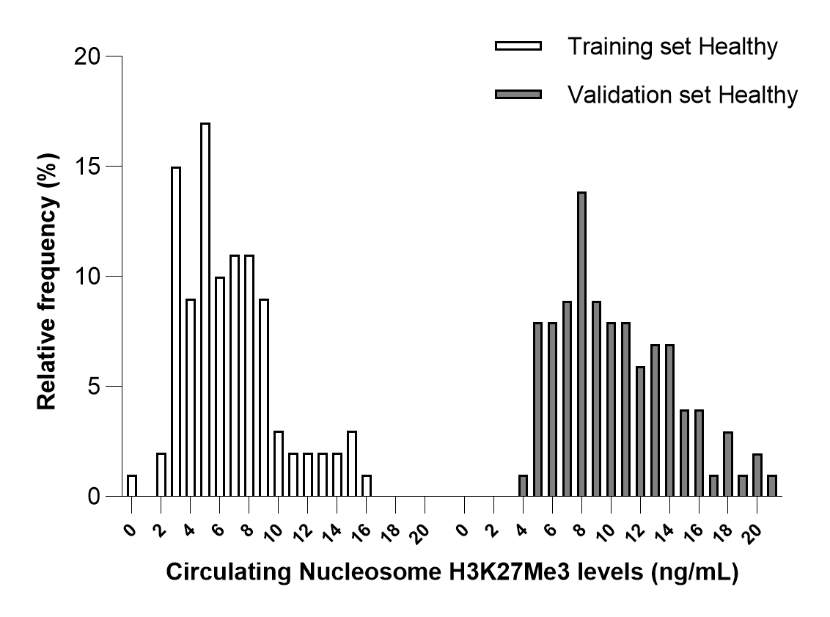
